# Design and rationale of the Longitudinal Evaluation of Norms and Networks Study (LENNS): A cluster-randomized trial assessing the impact of a norms-centric intervention on exclusive toilet use and maintenance in peri urban communities of Tamil Nadu

**DOI:** 10.1101/2020.06.26.20140830

**Authors:** Sania Ashraf, Cristina Bicchieri, Maryann G. Delea, Upasak Das, Kavita Chauhan, Jinyi Kuang, Peter McNally, Alexey Shpenev, Erik Thulin

## Abstract

**Introduction:** Inconsistent toilet usage is a continuing challenge in India. Despite the impact of social expectations on toilet usage, few programs and studies have developed theoretically grounded norms-centric behavior change interventions to increase toilet use in low-income settings. This protocol details the rationale and design of an ex-ante, parallel cluster-randomized trial evaluating the impact of a demand-side, norms-centric behavior change intervention on exclusive toilet use and maintenance in peri-urban Tamil Nadu, India.

**Methods and Analysis:** Following two years of formative research, we developed an evidence-based norm-centric behavior change intervention called *Nam Nalavazhvu* (Tamil for “Our wellbeing”). The multi-level intervention aims to shift collective beliefs by shifting empirical expectations or beliefs about other relevant people’s sanitation practices. It also provides action-oriented information to aid individuals to set goals and overcome barriers to build, consistently use and maintain their toilets. This trial includes 76 wards in Pudukkottai and Karur districts, where half were randomly assigned to receive the intervention and the remaining serve as counterfactuals. During baseline and endline (conducted one year after the initiation of intervention) assessments, we collect relevant data and compare results between study arms to determine the impacts of the *Nam Nalavazhvu* intervention on sanitation-related behavioral, health, wellbeing outcomes and potential moderators. This study is powered to detect differences in the prevalence of exclusive toilet use between study arms. We will also conduct a process evaluation to understand the extent to which the intervention was implemented, as designed.

**Ethics and Dissemination:** The study protocol has been reviewed and approved by the ethics board at the University of Pennsylvania, USA and the Catalyst Foundation, India. Research findings will be disseminated through open access peer reviewed publications and presentations to stakeholders, government officials and conferences.

**Trial registration:** NCT04269824.

**Strengths and limitations of this study:**

- This ex-ante, parallel cluster randomized trial assesses the impact of a norm-centric behavior change intervention strategy to improve sanitation practices. These behavior change techniques are novel to the sanitation sector but has been effective in changing a variety of behaviors, such as water use, drinking behavior, and energy consumption.
- The study outcomes include health, wellbeing outcomes, and a careful assessment of changes in social beliefs, expectations, and social determinants of collective sanitation behaviors.
- This study is being rolled out during the ongoing COVID-19 pandemic. This can potentially impact the effectiveness of this intervention package that uses community and network-based group activities. However, through a detailed process monitoring and evaluation we will be able to assess the impact on delivery, and subsequent behavior change in this unique setting.

## Introduction

### Study rationale

Low toilet use enables environmental contamination and contributes to poor health, wellbeing, and safety globally ^1-3^. In the past decade, national sanitation programs that aimed to curb persistent open defecation practices in India, including the Swachh Bharat Mission (SBM), significantly increased coverage of individual and shared toilets and promoted their use^4,5^. Despite increased access to toilets, complex sociocultural norms, technological and financial barriers prevented exclusive toilet use among low-income Indian communities ^6–8^. Recent behavior change interventions designed to promote toilet use in rural India have yielded, on average, a 5% increase in reported use amongst toilet owners, which is comparable to results generated by SBM.^9–11^. Sustaining exclusive toilet use among all household members is a national priority for the current Swachh Bharat Mission 2.0, also known as the Open Defecation Free (ODF)-plus scheme^12,13^. This is in line with the United Nations’ Sustainable Development Goal 6.2, which calls for the ‘achieve access to adequate and equitable sanitation and hygiene for all and end open defecation’ by 2030^14^.

Numerous studies have highlighted the importance of social beliefs and preferences on toilet construction and adoption^15–18^. Previous studies in India have specifically noted the role of social beliefs and perceptions in sanitation practices, including ownership and usage^6,7,10,19–21^. Social networks were also found to be relevant; a study in rural Karnataka showed that individuals were more likely to own toilets if their social contacts owned one22. Following the achievements of SBM, in a context of high toilet ownership and use, perceptions of others’ toilet ownership and (dis)approval might be particularly important motivators of toilet-related behavior change in India^10,19^.

Previous behavior change studies that assessed psychosocial determinants of toilet use and included norm-based messaging to promote toilet use in India were primarily guided by Community Led Total Sanitation (CLTS), the Risks, Attitudes, Norms, Abilities, and Self-regulation (RANAS) approach, or the Behavior Centric Design (BCD). All these approaches include norms as one driver of behavior^10,23–25^. No known study has used the “norms- diagnostic” approach detailed in the Social Norms Theory (SNT) to diagnose the social determinants of toilet use, which can then be leveraged to inform specific components of behavior change strategies^26^.

In our study, we aim to address several knowledge gaps in the norms-based literature with regards to sanitation behavior change. Our first objective was to diagnose the nature of collective patterns of toilet use in peri-urban communities of Tamil Nadu as a part of the formative research for Longitudinal Evaluation of Norms and Networks Study (LENNS)^28^. Such collective behaviors may or may not be socially conditional. Social conditionality means that the preference for adopting a specific behavior is driven by social expectations. Social expectations refer to a specific reference network, that we measure and may vary depending on the target behavior. These expectations can be empirical or normative (i.e., beliefs about what relevant others do or beliefs about what relevant others think one should do, respectively). In our formative research, we assessed the degree to which the preference for toilet use was conditional on these expectations to determine whether descriptive or social norms drive toilet use^26–27^. We found that toilet use was conditional on empirical, but not normative expectations, and therefore diagnosed toilet use in these communities as a descriptive norm^28^. This is consistent with other recent studies that reported empirical expectations as a significant psychosocial determinant for toilet ownership^10,16^. Based on these findings, we designed a theoretically-grounded, evidence-based behavior change intervention, and will evaluate its effectiveness on the uptake of exclusive toilet use and maintenance through this randomized trial.

Second, prior studies have not explored norms-based intervention techniques specifically designed to change empirical expectations of sanitation behaviors. An additional objective of this study is to test novel techniques and implementation approaches in low-income communities to change descriptive norms and downstream sanitation behaviors.

Third, there is only limited evidence on whether intervening upon empirical expectations of others’ sanitation behaviors can lead to the emergence of normative expectations of toilet use. Such a phenomenon is theoretically plausible, and if demonstrated through this study, our findings can lead to insights that can inform the design of behavior change strategies ^29,33^. Lastly, our study is based in peri-urban communities, which will add to the currently limited sanitation literature regarding interventions that benefit peri-urban populations ^30–32^.

## Objectives

This study protocol summarizes the rationale and methods of a cluster randomized trial (LENNS) that aims to evaluate the impact of a multi-level, demand-side behavior change intervention package called *Nam Nalavazhvu* on exclusive toilet use and maintenance.

The *Nam Nalavazhvu* intervention design leverages two years of formative research that found toilet use in peri urban Tamil Nadu is a descriptive norm (i.e., toilet is conditional on the belief that most relevant others are using a toilet). The intervention reflects a norm- and network-centric approach that will employ dynamic information dissemination to signal others’ sanitation practices while also addressing other barriers to the adoption of improved sanitation practices^28^.

The primary research aim of this study is to evaluate the impact of the *Nam Nalavazhvu* intervention on behavioral and health outcomes, specifically on:

- Exclusive toilet use among individuals aged five years and older (primary outcome) Secondary aims include assessing the impact of the intervention on:
- Coverage and access to improved toilet
- Maintenance of sanitation facilities
- Empirical expectations, normative expectations, and other behavioral antecedents
- Mental wellbeing of respondents
- Diarrheal and respiratory health of all household members

## Methods

### Study setting

The study is being conducted in 76 wards in peri-urban areas of Pudukkottai and Karur districts in Tamil Nadu, India (Figure 1). The unit of randomization for this study is the ward, the smallest administrative unit of a Town Panchayat (TP). According to the 2011 Census, these are urbanizing districts, where in Pudukkottai and Karur respectively, the residents were mainly agricultural laborers (31%, 34%), workers in industries (1.3%,1.2%) and other private businesses (16%, 43%). These districts are comprised primarily of Hindus (88%, 93%) and a minority of Muslims (7.1%, 5.1%). While both districts were declared open defecation free in October 2019, there were variations in toilet coverage and use across constituent wards, which we captured through informal conversations with officials.

**Figure 1:**
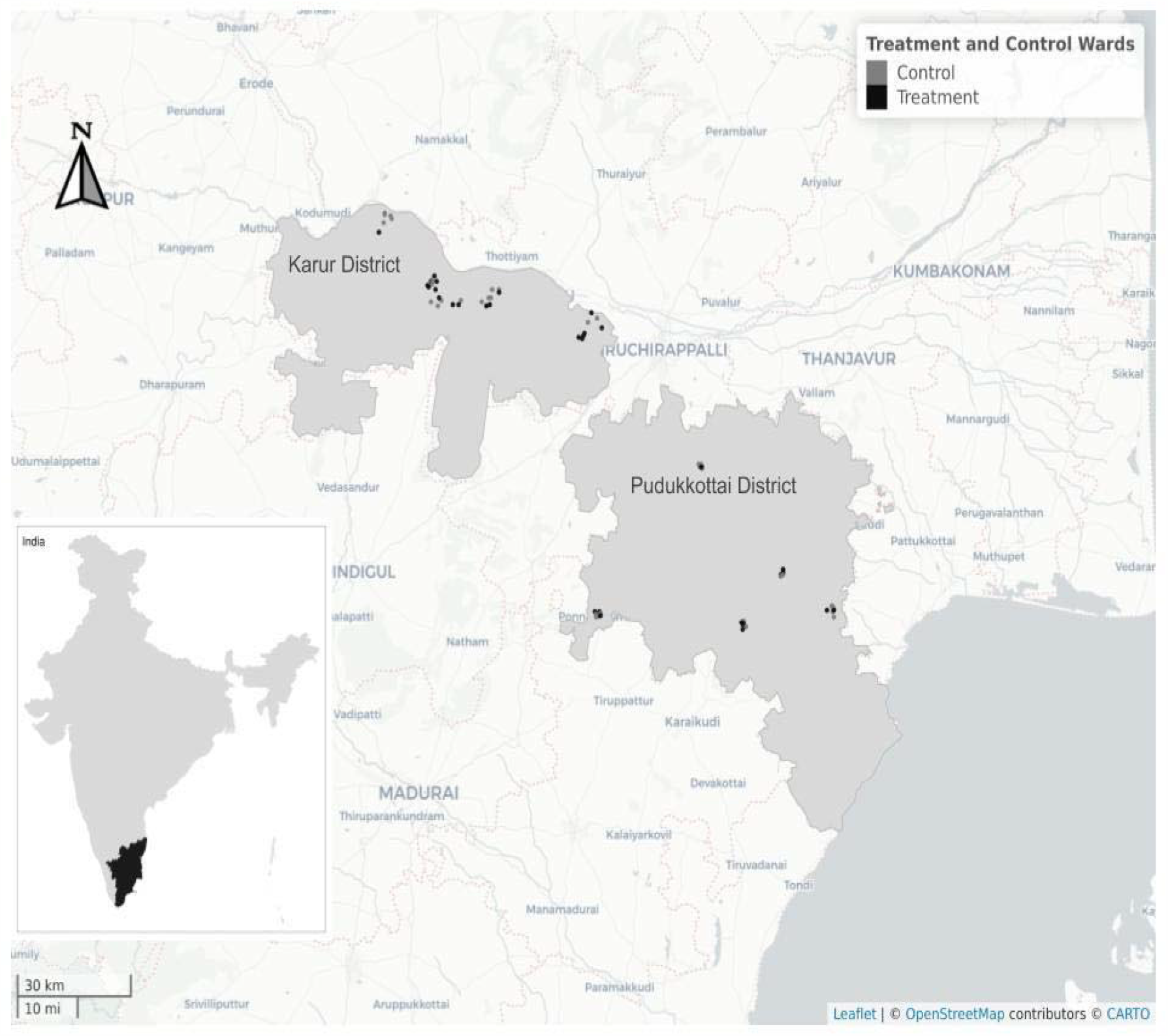
Study sites in Karur and Pudukkottai districts of Tamil Nadu, India.

### Study design

The LENNS trial is an ex-ante, parallel cluster randomized controlled trial (CRT). We will collect data relevant to our research questions in both study arms and compare results between intervention and counterfactual arms to determine the impact of the intervention on the primary and intermediate outcomes. This study is powered to detect differences in the prevalence of exclusive toilet use between study arms.

Clusters are defined as wards within purposively selected town panchayats. Half of the clusters were randomized to receive the *Nam Nalavazhvu* intervention, while the other half will not receive any active intervention as part of study. Prior to our enrollment of study participants, we created a buffer zone, minimally one ward in distance, between study clusters to minimize spillover. Since this study is being conducted in a context where the Indian government is actively implementing Swachh Bharat Mission – Urban (SBM), both study arms may be subject to government-led sanitation program activities.

The *Nam Nalavazhvu* intervention is further detailed in a following section. We co-designed this intervention with our implementation partner, a local NGO called Swasti (https://swasti.org/). Prior to the CRT, we conducted a three-month trial of improved practices (TIPs) to test, refine and revise our behavior change intervention activities and materials among a separate population in the same study districts (Fig 2). In the CRT, the intervention will be implemented for 12 months. A baseline and one-year follow up survey will be used to assess the impact of the intervention. We will also conduct a process evaluation to determine the extent to which the intervention was implemented, as designed, and identify successful pathways or barriers to the adoption of improved sanitation behaviors. Our process evaluation will assess fidelity of intervention implementation, reach, and contextual changes in community and household conditions that may facilitate improved behavioral adoption and/or outcomes. In addition, we will also conduct qualitative research with respondents and stakeholders to assess exposure and dose received of the intervention. Further, we aim to assess the extent of spillover in the control and the adjacent wards using mixed method research tools.

**Fig 2:**
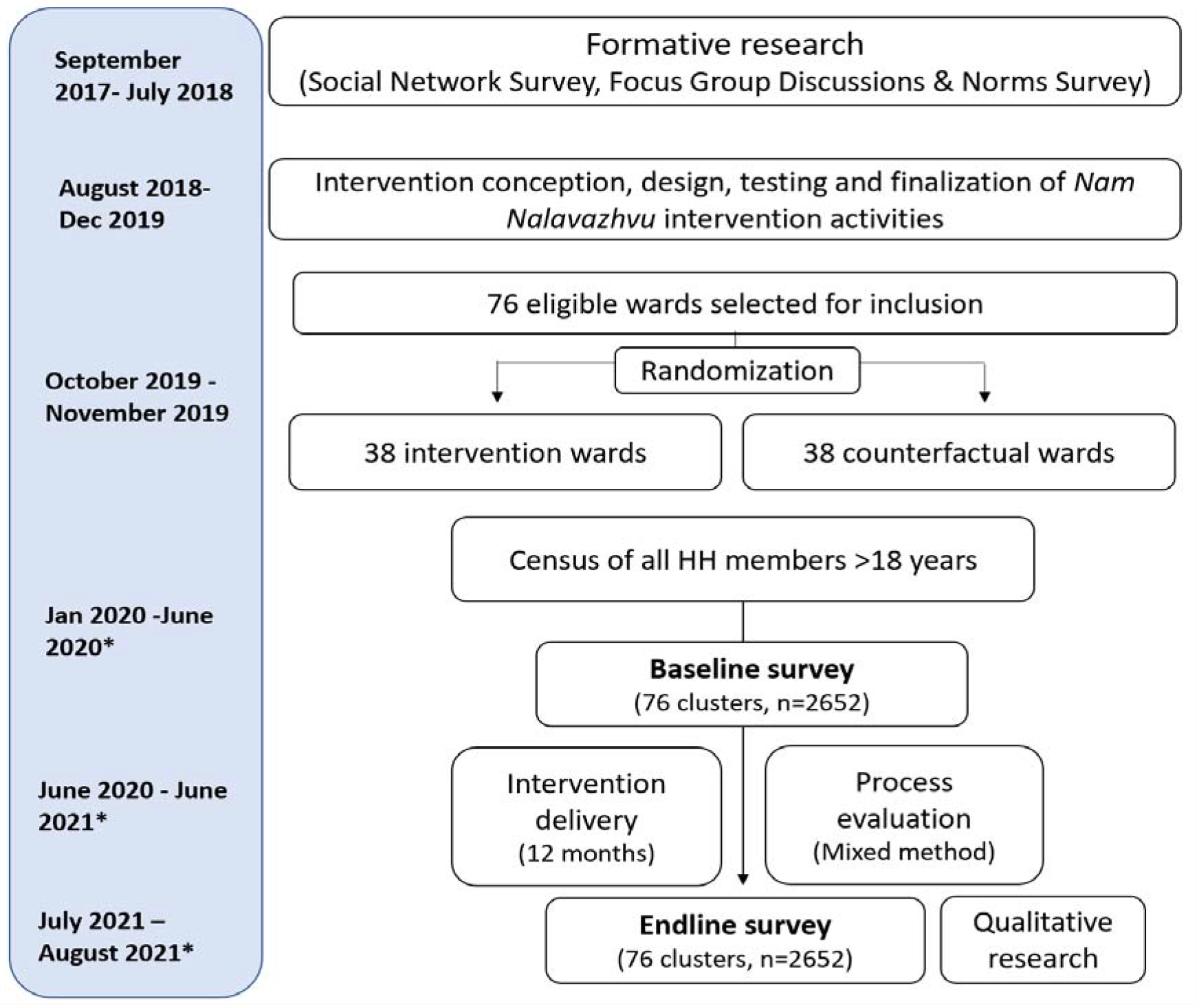
Summary of overall study design, including the timeline for impact evaluation to assess the effectiveness of the *Nam Nalavazhvu* intervention in Tamil Nadu, India *Timeline subject to change due to the COVID-19 related suspension in operations

### Description of intervention

*Nam Nalavazhvu* means ‘Our Wellbeing’ in Tamil and reflects a demand-side, norm-centric intervention designed to improve exclusive use of sanitation facilities for defecation purposes. Intervention activities focus on i) shifting empirical expectations by broadcasting improved sanitation behavior of relevant others in the community through activities at all levels (i.e. norm signaling) ii) capacity building and increased action knowledge through expanded social networks and access at the group level. It will not directly provide hardware or build toilets but address opportunity limitations. Details of the intervention design process, theoretical and behavioral framework will be published in separate forthcoming articles.

This multi-level intervention includes activities at the individual, household, group, and ward levels (Box 1).

#### Box 1

***Nam Nalavazhvu* intervention activities for peri urban area, Tamil Nadu**

At the ward level:

- Community mobilization and public commitment events
- Mass media broadcasting descriptive information
- Wall paintings promoting toilet ownership and use

At the group level:

- Peer learning sessions
- Descriptive norm messages delivered to social network members via community influencers

At the household level:

- Tailored counselling sessions with household members
- Visual signals of improved sanitation practices

At the individual level, in concurrence with household counselling sessions:

- Personalized advice, including information regarding similar others’ practices

### Dynamic descriptive information dissemination and capacity building

To update empirical expectations regarding others’ improved sanitation practices, we will periodically collect information about sanitation practices from all households in study clusters. We will then disseminate related descriptive norm information during community events, household visits, and text messages from their peers. Action knowledge and social connections between ward members, will be increased by disseminating contact information of neighbors, outreach workers and similar others in the ward who have adopted improved sanitation practices. Details about available financial schemes, sanitation markets and masons will be provided, and the outreach workers will assist in setting goals and taking steps to make progress towards them. Details of the activities are as follows:

#### Ward Level

Prior to the intervention delivery, ward outreach workers (WOWs) meet local stakeholders to conduct a social and resource mapping to engage them, foster support for the project goals and identify influencers who can be leveraged during the activities. Preliminary meetings will be conducted with these influencers (e.g., teachers, religious leaders, self-help group coordinators) to mobilize them. WOWs will also leave their contact cards to establish communication channels.

##### Roving audio announcements

Automobiles with loudspeakers will announce the launch of the program and invite community members to group sessions and community events. The audio content will include customized jingles to promote toilet use and later disseminate ward level data regarding people’s actual and intended sanitation practices.

**Wall paintings** (6*4 feet) will be used in at least 4 public locations per ward. Permissions will be secured from required private or government officials to paint the publicly visible wall. The motto, tagline and norm-centric images will be painted to encourage others to “Join the proud toilet owners”.

##### Community Mobilization and Commitment Events

Participants will include influential political and community leaders to applaud exclusive toilet users and promote improved sanitation practices. Messages will highlight the change happening in their communities, the benefits of using a toilet for families, and facilitate a public commitment to exclusive toilet use. These messages will reinforce those residents hear through other group and household activities.

#### Group Level

##### Peer learning session

Six sex-segregated groups will be convened in each ward over one year to facilitate social networking between toilet users and non-toilet users. The WOW supervisor will facilitate these 60-90-minute sessions. He/she will use SBCC materials (story cards/video content) to facilitate norm-focused conversations and information sharing among group members on how to access sanitation markets, barrier identification, planning, and coping by discussing personal experiences, challenges, solutions regarding target sanitation practices. Peer-to-peer knowledge sharing will be encouraged.

##### Social media to reference networks

Influential community members who are active on social media will be recruited as volunteer promoters or advocates. We will invite them to use their social media groups to send key messages promoted by the project. The LENNS team will develop messages and monitor their deployment as per schedule. These advocates will also use their social groups to broadcast testimonials from toilet adopters, related community events and activities.

#### Household Level

**Household counselling visits** allow outreach workers to get information about the household’s potential for change. The outreach workers communicate ward-specific toilet use of similar households to motivate change. Flipbooks and story cards will be used to engage and motivate household members. The outreach worker will counsel them how to achieve their goals by mapping out the next steps using goal cards. These households will also be informed of their neighbors’ improved practices. WOWs will focus on those lagging behind to motivate them to change, signaling others like them who already have.

##### Visual signals of improved practices

Improved behaviors of neighbors are signaled using bright decals or stickers, placed in public view on the household wall. These aim to serve as a goal and a source of pride for the households who receive and display them.

The implementation team is hired and managed by Swasti. Investigators at the Center for Social Norms and Behavioral Dynamics (CSNBD) at the University of Pennsylvania provide training, technical input, and oversight to the implementation process. The main delivery agent is a ward outreach worker (WOW). They are residents of the ward with a minimum of 12 years of formal education. Field supervisors help organize and facilitate group and ward-level activities. In addition, the study includes influential members from the community to disseminate promotional and motivational messages to their social network.

Participation in these activities is voluntary. The audio announcements and visual cues or paintings in public spaces will be apparent during the intervention. Ward outreach workers will consider any requests to adjust the recipient’s level engagement in the activities to ensure consent and comfort of participants.

##### Public involvement

As mentioned in the intervention details, members of the public are used to disseminate promotional messages through social media, using decals, active participation in groups and community events. We also incorporated their feedback in improving intervention delivery techniques and platforms prior to the trial.

##### Eligibility criteria

We randomly selected five town panchayats from each district. Researchers visited several wards with local Swasti officials to ensure they reflected the generalized peri-urban setting. To identify potential study wards within these TPs, we met local executive officers to assess ward maps. We used the following exclusion criteria:

- Commercialized wards with few residential households
- Urbanized wards with known high or complete coverage of improved toilet access according to TP official records
- Wards which bordered two or more adjoining wards, to reduce spillover

Our sampling frame excluded TPs where we piloted our interventions (n=3), or those where we did not receive permission to proceed due to political concerns (n=1).

The unit of randomization for this CRT is the ward. The demarcation of the ward is derived from the 2011 census conducted by the Ministry of Home Affairs, Government of India^34^. Trained field teams generated ward maps to delineate the ward boundaries, confirmed it with residents, and ensured a buffer zone. We also engaged ward-level stakeholders to assist in the mapping process and gain access to communities.

All ward residents from intervention clusters were eligible to participate in the intervention activities. For data collection, field workers surveyed randomly selected household members who were 18 years or above, planned to reside in that household continuously for the next year and were willing and able to participate.

### Selection and assignment of interventions

Following the baseline data collection, a co-investigator (UD) randomly assigned the wards to counterfactual and intervention arms in a 1:1 ratio, using a computer-generated randomization sequence. The randomization is geographically pair-matched within each town panchayat (Table S1). Balancing the study arms across geography will allow us to adjust for spatially clustered features or events that may be associated with our outcome. Given the nature of the intervention, the study investigators and the participants will not be masked to the intervention assignment. The data collection team will be masked to the treatment assignment.

### Recruitment

Trained enumerators conducted a listing exercise to generate a sampling frame of eligible individuals within the ward. Enumerators approached the household and asked the selected individual if they would consent to participate in the study. If the selected individual was absent or unavailable, up to three repeat visits were made to enroll them. If unsuccessful, the enumerators approached the next randomly selected individual from that cluster. Following randomization, households in intervention wards will be enrolled following an oral consent process.

The individuals enrolled in qualitative studies will be purposively selected from intervention wards to address relevant research questions. Those sampled from the counterfactual wards will allow us to investigate spillover of the *Nam Nalavazhvu* intervention.

### Sample size

This study is powered to detect differences in the prevalence of exclusive toilet use between study arms. Measuring exclusive use is problematic due to recall and desirability bias. As a result, during our formative research phase, we asked individuals about their defecation place the last time they needed to defecate during two rounds of data collection conducted 8 months apart. We incorporated the correlation between these two measures in our sample size calculation. We powered our study at 80% based on the prevalence of reported toilet use during last defecation in peri urban Tamil Nadu (estimated at 64.4% in 2018) and assumed a 10-percentage-point improvement as the minimum important effect. In order to detect such a minimal effect in exclusive toilet use (given observed intra-cluster correlation of 11% and correlation of last use with the one measured in the fall of 2017 of 47.5%), we estimated a requirement of 76 clusters (38 clusters per arm). We require 30 individuals per cluster. Assuming 10% loss to follow up, we engaged 34 individuals per cluster for a total of 2280 individuals. Since we collected household level toilet usage data, we have data for more individuals beyond the actual respondent.

### Study outcomes and measures

We will evaluate these outcomes at baseline and at endline, one-year after the intervention implementation using verbal surveys, in both intervention and counterfactual clusters. We will characterize sanitation facilities using standardized categorizations through direct observations.

### Primary outcome

The primary outcome for this trial includes:

1. Proportion of households where all members (18+ years) exclusively use a toilet every time they defecate. We will combine responses to several self-reported toilet use behaviors to determine exclusive toilet use in the past two days. We will also observe toilets to check for signs of use. (See S2)

### Secondary outcomes

Secondary outcomes of interest include:

1. Presence and access to improved toilets will be assessed using standard questions. Spot observations of toilets will be used to assess maintenance, functionality, and recent use
2. Mental wellbeing will be measured using the World Health Organization-Five Well-Being Index (WHO-5)
3. Diarrheal disease for all household members will be measured using the WHO definition of three or more loose stools in a 24-hour period, with or without the presence of blood
4. Respiratory illness for all household members will be measured using reported cough and/or difficulty breathing/shortness of breath according to the WHO’s Integrated Management of Childhood illness (IMCI)
5. Intermediate behavioral antecedents such as empirical expectations (i.e., what other people do), normative expectations (i.e., what other people think one should do) of prevalence of toilet ownership, exclusive use and maintenance using tested indicators (See S2)

## Data collection

Randomly selected individuals were enrolled in the impact evaluation. They responded to a baseline survey and will be approached to complete a one year follow up survey administered by trained enumerators. The enumerators will be masked to the intervention assignment. However, given the nature of the intervention, they might observe *Nam Nalavazhvu* intervention products in the household during the follow up survey.

We will consider respondents lost to follow up if: 1) they refuse to participate in the follow up survey, 2) if they have relocated elsewhere outside the intervention ward, 3) the field team is unable to reach them despite three attempts during data collection.

CSNBD researchers will work with trainers to conduct a 10-day training session prior to each survey round. The training sessions will be conducted in Tamil. Prior to the baseline survey, the instrument was administered to respondents similar to the target group to ensure comprehension. We incorporated feedback to clarify language, framing, answer choices and administration of the survey. The survey data will be collected using personal handheld devices. These electronic surveys were tested to address issues with data capture, skip patterns, and validity checks for each item in a pilot study. Quality assurance steps will be taken to improve data accuracy including regular field-level data checks and dual data capture of objective measures from a subset of households by field supervisors. Researchers from CSNBD will also visit the sites randomly to assess the situation pertaining to the survey. Weekly phone meetings will be conducted with the data collection agency to ensure quality of data.

The implementation partner will collect routine monitoring data to capture information about intervention fidelity and exposure. We will take steps to ensure that they do not involve participants enrolled in the impact evaluation to reduce participant fatigue.

For process documentation, we will conduct three rounds of qualitative data collection as part of the process documentation for this study. These will be at 3 months, 6 months and 1 year after the start of the intervention and assess participant response, acceptability, and the beliefs about the improved sanitation behavior among respondents in the intervention and counterfactual groups. Trained qualitative researchers will conduct in-depth interviews (IDI) and/or focus group discussions (FGD) with purposely selected respondents in intervention clusters to assess their interaction and experience with the *Nam Nalavazhvu* intervention. They will use semi-structured questionnaires, memos and note taking to record observations and may record interviews as required. Verbal consent will be taken before every data collection activity except observations made in public spaces.

## Data management

All survey data will be transmitted through secured servers and stored in in password protected folders in the Penn Box. To protect confidentiality, all subjects will be deidentified for analysis. Data will only be accessible to the Penn faculty, staff, and data management personnel.

## Statistical analyses

We will use intention to treat (ITT) analyses to assess the difference in specific outcomes between study arms after exposure to one year of the *Nam Nalavazhvu* intervention. For most of the outcomes, we will use a log-binomial regression model to assess prevalence ratios of post-intervention sanitation-related outcomes across intervention groups. We will consider adjusting for variables that were imbalanced between the groups at baseline in adjusted models. We will also use generalized estimating equations (GEE) with robust standard errors to account for the clustering of observations within each cluster (ward). We will use post-estimation commands to estimate and report the average marginal effects. We will not adjust p values based on multiple comparisons.

In additional analyses, we will use appropriate multivariate models to assess the impact of the interventions on secondary outcomes. Both unadjusted and adjusted effect estimates will be reported for all outcomes. Following the process evaluation findings, if fidelity or intervention quality varied considerably in the trial, we will consider a per-protocol or other appropriate analyses to assess the impact of the *Nam Nalavazhvu* interventions on our outcomes of interest. The analyses will be conducted by the scientific team including statistical software including R and Stata.

## Ethics and dissemination

The ethics review board at the University of Pennsylvania (833854) and the Catalyst Foundation, India reviewed and approved this research protocol. The trial is registered with clinicaltrials.gov: NCT04269824. All amendments and protocol modifications will be updated there. All enrolled study communities provided verbal consent to enroll in the study. Surveyed individuals will provide written informed consent. This consent process was conducted in the local language, Tamil. Participants will receive messages that may encourage them to improve their sanitation conditions or practices. Our assessment is that the benefits to study participation outweigh the minimal risks. Deidentified data will be used during analysis.

Research findings will be disseminated through presentations at conferences and submitted to peer reviewed journals for open access. Our results will be shared with relevant local stakeholders through community-based meetings in participating wards through presentations made to the district and state level officials in Tamil Nadu.

## Data monitoring, reporting harms, and auditing

The research team has text message groups and weekly calls with the implementation partner, to discuss progress, issues from the field including adverse events so that prompt action can be taken. No harm is anticipated to the intervention recipients in this study. There are no plans for a data monitoring committee or audits for this trial.

## Discussion

This study will evaluate the effectiveness of the *Nam Nalavazhvu* intervention, a demand-side, descriptive norms- and network-centric intervention approach that aims to shifting empirical expectations on targeted sanitation behaviors. Our behavior change communication approach employs dynamic signaling (i.e., dissemination of descriptive information regarding others’ actual and/or intended improved sanitation practices), and reflects a strategy that is novel to the sanitation sector, yet has been effective in changing a variety of behaviors, such as water use, drinking behavior, and energy consumption ^35–38^. Evaluating the impact of such an approach may have widespread implications for policy and practice for sanitation programming in India and beyond if the intervention proves effective in improving sanitation behaviors via changing people’s empirical expectations. Our plan to also track normative expectations will also allow us to determine whether an intervention focused on shifting of empirical expectations has spillover effects on normative expectations.

Evidence from this study will address knowledge gaps regarding the application and effectiveness of a “norms-diagnostic” approach in the design of behavior change strategies that intervene upon the social determinants of collective sanitation behaviors. The intervention uses outreach workers and social media users to deploy most of its messages. Understanding the transmission of messages during household visits, peer learning sessions and text messages will inform recommendations on the feasibility and effectiveness of using these platforms for norms-centric interventions. Insights generated in this study may contribute to the tools available to address descriptive norms for community-based interventions, generally.

Limitations of this CRT include the use of wards as clusters in peri-urban communities. Although these are the smallest geographic operational units, some boundaries in specific districts were redrawn following the start of the intervention, leading to concerns about spillovers across buffer areas. Two critical country specific incidences are impacting the implementation of the interventions. One is the highly contentious Citizens Amendment Act (CAA) passed in Dec 2019, that led to nationwide protests in India. In our study it led to refusals by households in predominantly Muslim wards, who resisted participating in any study that include survey-based instruments. Secondly, the ongoing COVID-19 pandemic has led to considerable interruption in the implementation of group-level intervention activities. These aspects are being assessed through our process evaluation and will inform the interpretation of the results from this CRT.

## Data Availability

Data will be available with the publication of the main outcomes paper from this trial at https://osf.io/bkm9n/. Socio-behavior change communication materials will be shared as supplementary materials in peer-reviewed publications in open access journals.

## Acknowledgement

We sincerely acknowledge the substantial insights and efforts of K. Jeyaganesh, Rajesh Kanna, Johnson Thangaraj, Raja Rethinam and the experienced team implementing this study at Swasti, Catalyst Management Services (CMS). New concepts, Delhi helped us design our socio-behavioral change communication materials following multiple rounds of field tests. We are also grateful for the time of our outreach workers and the respondents who provided feedback during the development of the intervention activities and messages.

## Availability of data and materials

Data will be available with the publication of the main outcomes paper from this trial at https://osf.io/bkm9n/. Socio-behavior change communication materials will be shared as supplementary materials in peer reviewed publications in open access journals.

## Author contributions

CB is the principal investigator for this study. She has developed the social norms theory (SNT) and the measurements that has driven the intervention design for this CRT. SA, MGD, AS, UD and CB designed this study. CB, MGD, ET and SA contributed substantially to the design of the multi-level behavior change intervention. SA, MGD, JK, UD and KC contributed to the testing and implementation of the intervention. CB, SA, MGD, AS, UD, PM and JK contributed to the development of data collection tools. AS, UD, SA, and KC supervised the data collection. SA wrote the first draft of the manuscript. All authors reviewed and contributed to the final manuscript. All authors read and approved the final manuscript.

